# Clinical consequences of a polygenic predisposition to benign lower white blood cell counts

**DOI:** 10.1101/2023.08.20.23294331

**Authors:** Jonathan D. Mosley, John P. Shelley, Alyson L. Dickson, Jacy Zanussi, Laura L. Daniel, Neil S. Zheng, Lisa Bastarache, Wei-Qi Wei, Mingjian Shi, Gail P. Jarvik, Elisabeth A. Rosenthal, Atlas Khan, Alborz Sherafati, Iftikhar J. Kullo, Theresa L. Walunas, Joe Glessner, Hakon Hakonarson, Nancy J. Cox, Dan M. Roden, Stephan G. Frangakis, Brett Vanderwerff, C. Michael Stein, Sara L. Van Driest, Scott C. Borinstein, Xiao-Ou Shu, Matthew Zawistowski, Cecilia P. Chung, Vivian K. Kawai

**Affiliations:** Department of Medicine, Vanderbilt University Medical Center, Nashville, TN, USA; Department of Biomedical Informatics, Vanderbilt University Medical Center, Nashville, TN, USA; Yale School of Medicine, New Haven, CT, USA; Department of Genome Sciences, University of Washington Medical Center, Seattle WA, USA; Department of Medicine (Medical Genetics), University of Washington Medical Center, Seattle WA, USA; Division of Nephrology, Dept of Medicine, Vagelos College of Physicians & Surgeons, Columbia University, New York, NY; Department of Cardiovascular Medicine, Mayo Clinic, Rochester MN USA2; Department of Medicine, Northwestern University Feinberg School of Medicine, Chicago, IL, USA; Department of Pediatrics, Children’s Hospital of Philadelphia, Philadelphia, PA, USA; Department of Pediatrics, Perelman School of Medicine, University of Pennsylvania, Philadelphia, PA, USA; Department of Pharmacology, Vanderbilt University Medical Center, Nashville, TN, USA; Department of Anesthesiology, University of Michigan Medical School, Ann Arbor, Michigan, USA; Department of Biostatistics, Center for Statistical Genetics, University of Michigan, Ann Arbor, Michigan, USA; Department of Pediatrics, Vanderbilt University Medical Center, Nashville, TN, USA; Vanderbilt-Ingram Cancer Center, Vanderbilt University School of Medicine, Nashville, TN, USA

**Keywords:** White blood cell count, polygenic score, bone marrow biopsy, taxane, azathioprine, pharmacogenomics

## Abstract

Polygenic variation unrelated to disease contributes to interindividual variation in baseline white blood cell (WBC) counts, but its clinical significance is undefined. We investigated the clinical consequences of a genetic predisposition toward lower WBC counts among 89,559 biobank participants from tertiary care centers using a polygenic score for WBC count (PGS_WBC_) comprising single nucleotide polymorphisms not associated with disease. A predisposition to lower WBC counts was associated with a decreased risk of identifying pathology on a bone marrow biopsy performed for a low WBC count (odds-ratio=0.55 per standard deviation increase in PGS_WBC_ [95%CI, 0.30 - 0.94], p=0.04), an increased risk of leukopenia (a low WBC count) when treated with a chemotherapeutic (n=1,724, hazard ratio [HR]=0.78 [0.69 - 0.88], p=4.0×10^-5^) or immunosuppressant (n=354, HR=0.61 [0.38 – 0.99], p=0.04). A predisposition to benign lower WBC counts was associated with an increased risk of discontinuing azathioprine treatment (n=1,466, HR=0.62 [0.44 - 0.87], p=0.006). Collectively, these findings suggest that a WBC count polygenic score identifies individuals who are susceptible to escalations or alterations in clinical care that may be harmful or of little benefit.

## INTRODUCTION

White blood cell (WBC) counts (the number of WBCs present within a volume of blood) are routinely measured in clinical settings to survey health, ascertain for drug toxicities and identify causes of illness. The counts are evaluated with respect to a reference range, the interval of values expected in a healthy population,^1,2^ and a measurement that falls outside of the interval may prompt investigations to exclude conditions such as infections, diseases of the bone marrow, autoimmune disease, and toxicities due to medications such as chemotherapeutics and immunosuppressants.^3^ A low WBC count may also prompt clinical action due to concerns that an individual may have an immunodeficiency that could limit an effective response to infections.^4^

A genetic predisposition toward benign lower WBC counts can impact clinical care. For instance, the rs2814778-CC genotype is common among individuals of African ancestry and is associated with lower WBC counts in the absence of underlying disease.^5,6^ Carriers of this genotype are more likely to undergo diagnostic investigations, including a bone marrow biopsy, and to have medications stopped due to concerns for toxicity.^7–10^ These actions are driven, in part, by the use of WBC count reference ranges that are not calibrated to this genotype.^11,12^ While the rs2814778-CC genotype variant is not prevalent among European ancestry (EA) populations, numerous common single nucleotide polymorphisms (SNPs) associated with WBC count variation have been identified in this group.^13,14^ Whether a polygenic predisposition toward benign lower WBC counts could have similar clinical consequences in this population is unknown.

We constructed a polygenic score for WBC counts (PGS_WBC_) which measures the burden of SNPs associated with WBC count an individual carries. SNPs located near loci associated with clinically significant diseases within the differential diagnosis of a low WBC count were excluded to ensure that the PGS_WBC_ measures benign WBC count variation. We examine a diverse range of clinical outcomes and settings to define the consequences of a polygenic predisposition to benign lower WBC counts.

## RESULTS

### Development and validation of a benign PGS_WBC_

We developed a polygenic risk score for WBC count (PGS_WBC_) using SNP weightings derived from a large genome-wide association study (GWAS) of WBC counts.^15^ A linkage-disequilibrium reduced (r-square<0.01) set of independent SNPs was selected (p<5×10^-6^, minor allele frequency=0.01, imputation r-square≥0.7). To ensure that SNPs associated with clinically significant diseases were not included in the PGS_WBC_, SNPs located in the major histocompatibility complex (MHC) genomic region, which is associated with multiple auto-immune diseases,^16^ were excluded as well as SNPs located near loci previously associated with hematological malignancies and systemic lupus erythematosus^17^ (see methods for full details). After exclusions, there were 1,739 WBC-associated SNPs in the PGS_WBC_. The PGS_WBC_ was normalized to have a mean of 0 and a standard deviation of 1, and a lower PGS_WBC_ value reflects a polygenic predisposition to lower WBC counts. Thus, an inverse association between the PGS_WBC_ and an outcome indicates that a predisposition to lower counts increases the risk of the outcome.

To verify that the PGS_WBC_ did not associate with clinically significant diseases that are in the differential diagnosis of a low WBC count, we tested for associations between the PGS_WBC_ and hematological (n=15) and auto-immune diagnoses (n=22) that were prevalent among 71,078 European Ancestry participants from BioVU, a DNA biobank linked to a de-identified electronic health record. There were no significant inverse associations (FDR p>0.05) with these diagnoses (**Supplementary Figure 1** and **Supplementary Table 1**).

We examined associations between the PGS_WBC_ a diverse range of clinical outcomes measured in different clinical settings (**Figure 1**).

**Figure 1:**
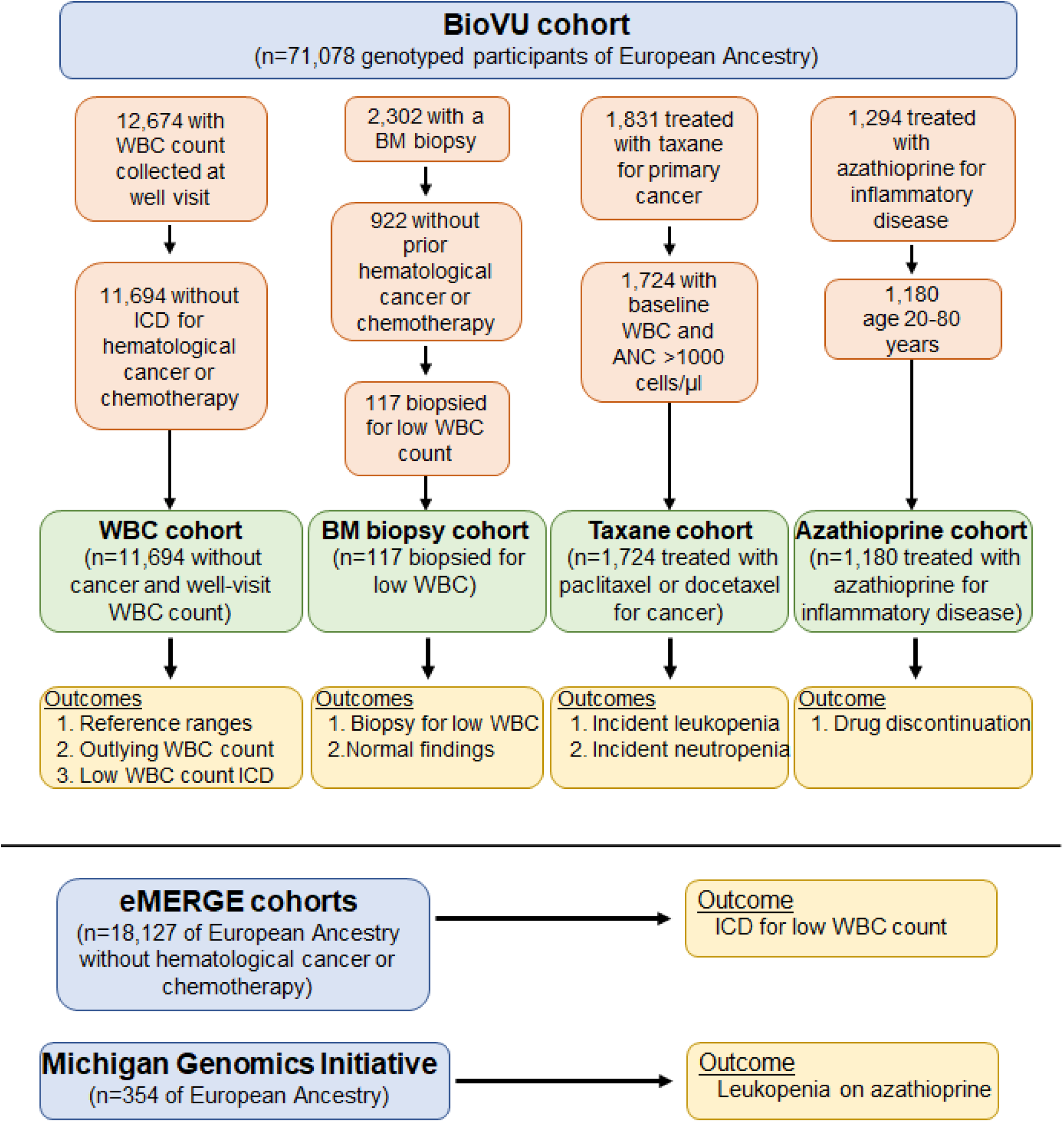
Overview of the study populations and analyses. WBC=white blood cell; BM=bone marrow; ANC=absolute neutrophil count.

### Association between the PGS_WBC_ and low measured WBC and markers of clinical activity

To define the relationship between the PGS_WBC_ and measured WBC counts, we identified 11,694 BioVU participants (6,931 females [59%]; mean age 57 [s.d. 17] years), without a hematological malignancy and who had 1 or more WBC count measurements collected in a primary care setting during a routine health maintenance exam. The PGS_WBC_ was positively correlated (partial correlation=0.29) with measured WBC counts (**Figure 2a**). WBC measurements are paired with a reference range specific to the clinical assay. There were 623 participants who had at least one WBC count that fell below the lower reference range (i.e. a value that would be designated as a clinical outlier). The PGS_WBC_ value was inversely associated with having a WBC count fell below the reference range (odds-ratio [OR]=0.57 [95% CI: 0.52 – 0.62]) per s.d. increase in the PGS_WBC_, p<2×10^-16^) (**Figure 2b**).

**Figure 2:**
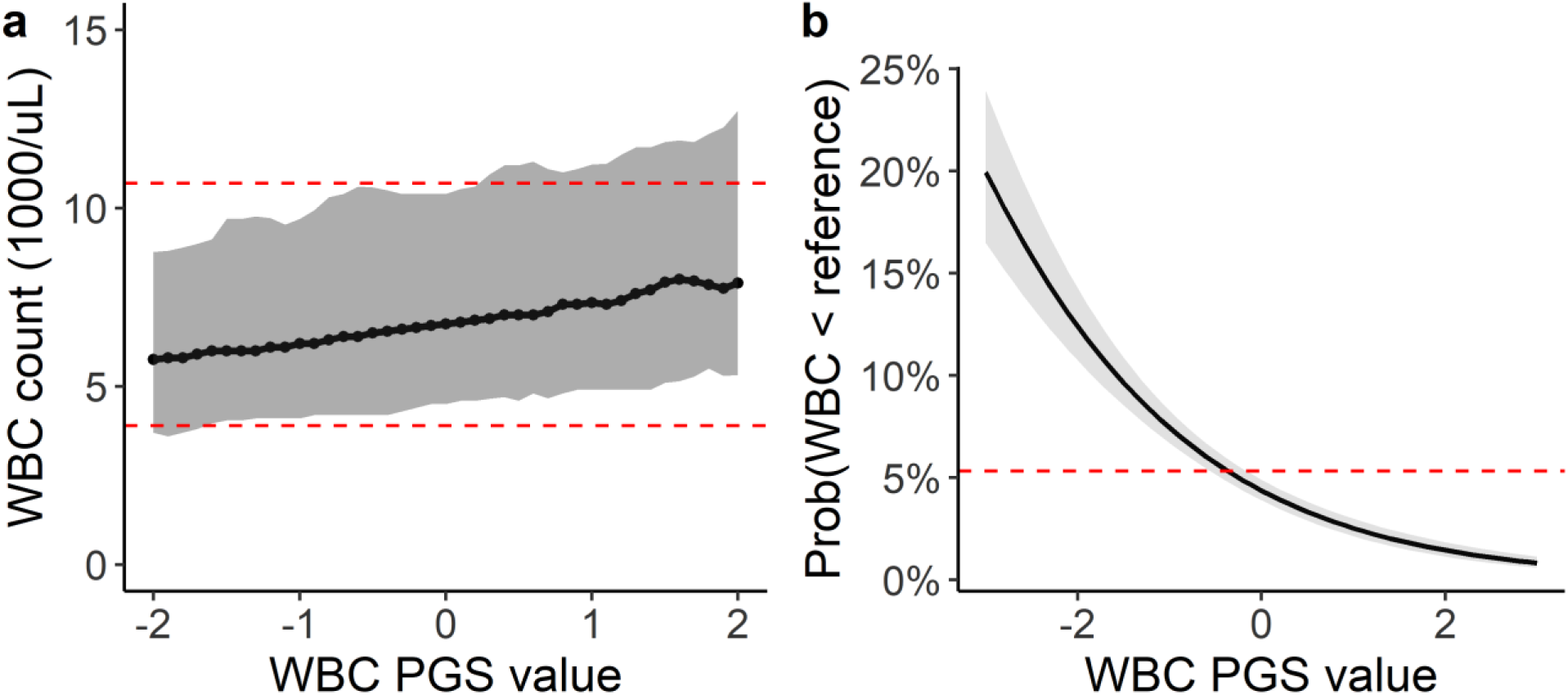
PGS associations with measured WBC counts among 11,694 BioVU participants. (**a**) Ranges of observed WBC counts by PGS_WBC_ value. Ranges summarize WBC counts within sequential windows (+/- 0.2 s.d.) across the range of the PGS_WBC_. The dark line is the median value and the grey ribbon represents the 5^th^ to 95^th^ percentiles of the range. The dashed red lines denote the upper and lower clinical reference ranges for the clinical assay used to measure the WBC count. (**b**) Predicted probability of having a WBC count that falls below the lower clinical reference value across range of PGS_WBC_ values. Probabilities are based on a logistic regression model adjusted for age and sex. The dashed red line is the average probability for the entire population.

ICD codes link clinical and billing activity to a patient’s underlying disorder.^18^ Having an ICD code for a low WBC count indicates that a diagnosis of low WBC count was made by a provider. There were 379 (3.2%) participants assigned an ICD code for a diagnosis of low WBC count. A lower PGS_WBC_ was significantly associated with having an ICD code in BioVU (OR=0.62 [0.56 – 0.69], p<2×10^-16^). A similar association was seen in an independent set of 18,217 participants from the eMERGE network who did not have a history of a hematological malignancy, where 256 (1.4%) participants had an ICD code for a low WBC count (OR=0.74 [0.67 - 0.82], p=2.0×10^-9^).

In sum, a polygenic predisposition toward lower WBC counts was associated with lower WBC counts, an increased likelihood of having a count below the clinical reference range and having an ICD code for a low count.

### Association between the PGS_WBC_ and bone marrow biopsies for low WBC

A bone marrow biopsy is an invasive procedure to determine whether a hematological abnormality, such as a low WBC count, is due to underlying disease in the bone marrow. Biopsy reports include a clinical indication, which lists the clinical concerns of the hematologist that prompted the biopsy. It has been previously observed that a benign WBC-lowering genotype was associated with both the clinical indication and outcomes of a bone marrow biopsy among individuals of African ancestry.^19^ We examined whether the PGS_WBC_ behaved similarly.

Among 922 BioVU participants without a prior history of a hematological malignancy who underwent a bone marrow biopsy, there were 117 who underwent a first biopsy where the clinical indication noted a concern for a low WBC count. The PGS_WBC_ was associated with having a biopsy for this indication (OR=0.56 [0.45 - 0.68], p=1.8×10^-8^).

Among the 117 participants who underwent a biopsy for an indication that included a low WBC count, 64 (55%) were female and the mean age was 48 (25) years (**Supplementary Table 2**). Bone marrow pathology was identified in 35 (30%) biopsies and was more frequent among participants who had other hematological comorbidities (e.g. anemia or a low platelet count) in addition to a low WBC count. Those without pathology in their bone marrow had PGS_WBC_ that were more skewed toward the lower ranges, as compared to those with pathology (**Figure 3a**). The PGS_WBC_ was associated with a finding of no pathology, after adjusting for other hematological comorbidities (OR=0.55 [0.30 - 0.94], p=0.04) (**Figure 3b**).

**Figure 3:**
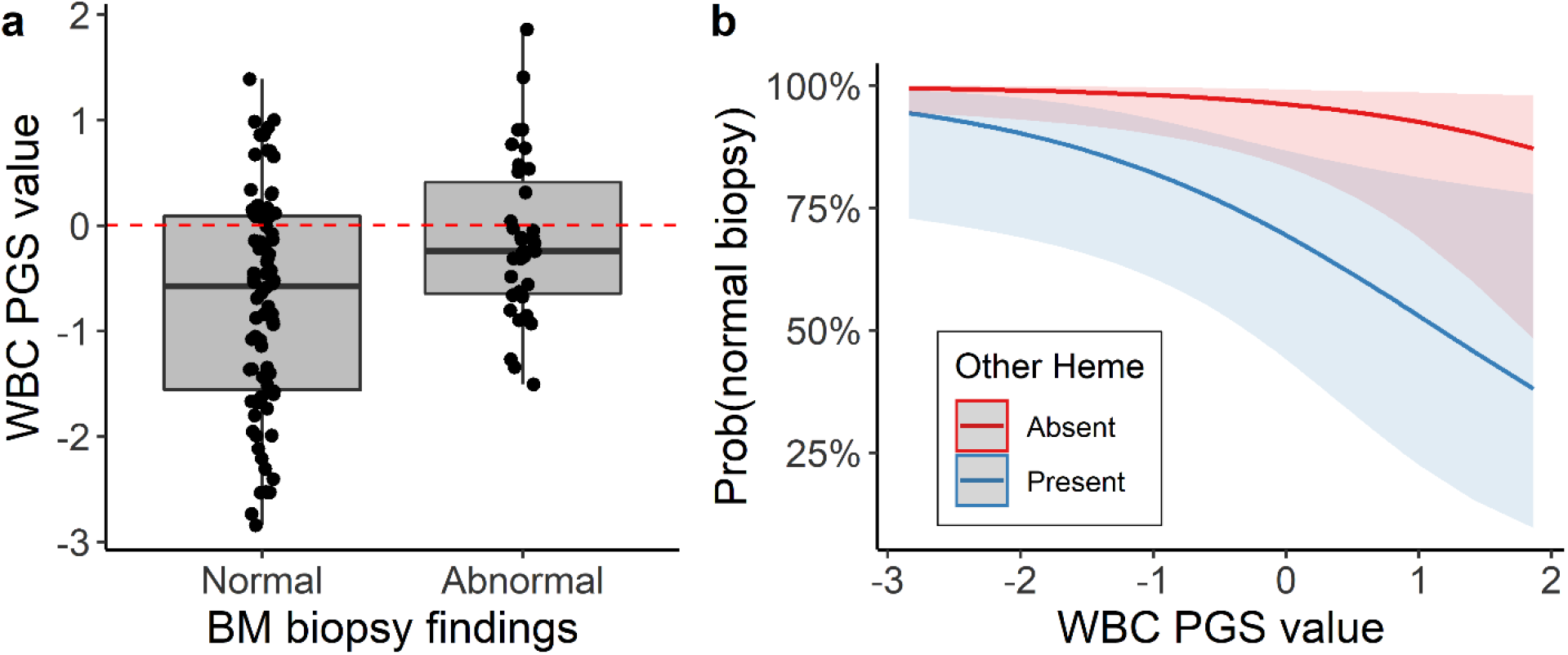
PGS associations with bone marrow biopsy outcomes. (**a**) Distribution of the PGS_WBC_ values among 117 BioVU participants who underwent a bone marrow biopsy for a clinical indication that included a low WBC count. Results are stratified by whether the biopsy identified pathology (abnormal, n=35) or not (normal, 82). (**b**) Predicted probabilities of a normal biopsy finding. Results are stratified by whether the indication for the biopsy include other hematological abnormalities in addition to a low WBC count. Probabilities are based on a multivariable logistic regression model, adjusted for age, sex and presence of a hematological abnormality.

In sum, a polygenic predisposition toward lower WBC counts was associated with an increased likelihood of having a bone marrow biopsy that was performed to investigate a low WBC count and a reduced likelihood of identifying pathology on the biopsy.

### Association between the PGS_WBC_ and drug-induced leukopenia

A benign predisposition to lower WBC counts has been shown to be associated with an increased risk of a low WBC count due to a medication (i.e. drug-induced leukopenia) as well as discontinuation of a medication due to a concern for low WBC counts among individuals of African ancestry with the rs2814778-CC genotype.^10,20^ We examined whether the PGS_WBC_ was similarly associated with drug side effects for two classes of drugs that can commonly cause drops in WBC counts: chemotherapeutics (antineoplastics) and immunosuppressants.

We first examined whether there was an association with ICD-based billing codes related to adverse effects from these classes of medications. Since these medications can be associated with a broad range of toxicities, we specifically examined whether the PGS_WBC_ was associated with having an ICD code for a toxicity due to chemotherapeutic or immunosuppressant medications entered on the same day that the provider wrote a clinical note that mentioned a low WBC count. In the BioVU population (n=71,078), there were 985 (1.4%) participants who met this case definition and the PGS_WBC_ was associated with this outcome (OR=0.92 [0.86 - 0.98], p=0.01) (**Figure 4a**). However, there was no association with the ICD codes in the absence of a clinical mention of a low WBC count (n=2273 cases, OR=1.01 [0.97 – 1.06], p=0.60).

**Figure 4:**
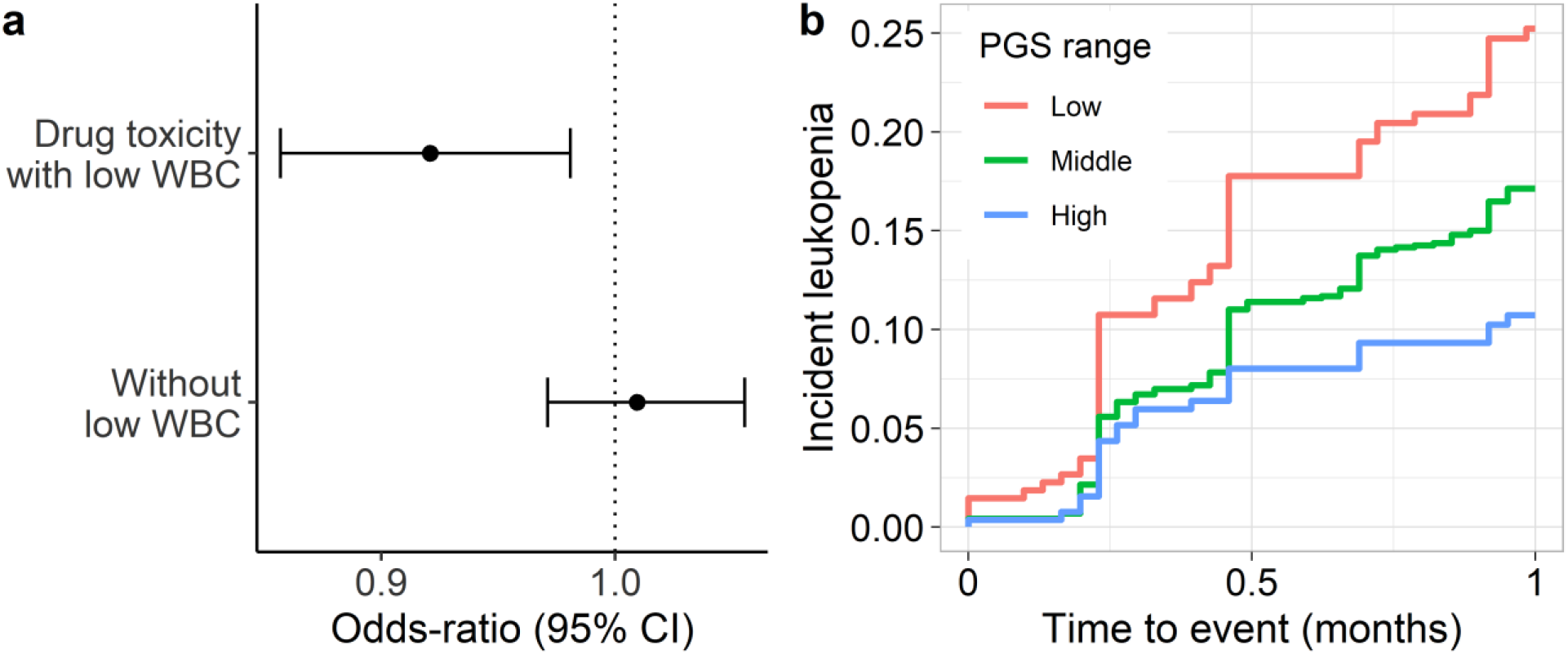
PGS associations with pharmacogenomic outcomes. (**a**) Odds-ratio of having an ICD-code for toxicity from an antineoplastic or immunosuppressive medication with (n=985) or without (n=2273) a clinical concern for a low WBC count among 71,078 BioVU participants. Odds-ratios are from a logistic regression model, adjusted for age, sex and principal components. (**b**) Kaplan-Meier plot for a WBC count<3,000 cells/ul (leukopenia) after initiating treatment with taxanes among 1,724 BioVU participants with cancer. The PGS_WBC_ strata are: Low (<1 s.d. below the mean), Middle (≥ -1 s.d. and ≤ 1 s.d.), High (>1 s.d).

To probe this association further, we identified 1,724 BioVU participants (917 [53%] female, 60 [12] years) who received treatment for cancer with the taxane class of chemotherapeutic medications^21^ (**Supplementary Table 3**). The mean baseline (pre-treatment) WBC count was 8,200 (s.d. 3,900) cells/µL and the PGS_WBC_ was significantly associated with baseline counts (log change per s.d. increase in the PGS_WBC_=0.062 [0.044 -0.080], p=3.9×10^-11^). In the first cycle of treatment, 266 (16%) participants developed an incident drug-induced leukopenia, defined as a WBC count<3,000 cells/uL. The PGS_WBC_ was significantly associated with this outcome after adjusting for treatment dose and duration (hazard ratio [HR]=0.78 [0.69 - 0.88], p=4.0×10^-5^) (**Figure 4b**). There was a similar association for the outcome of drug-induced neutropenia (absolute neutrophil count<1,500 cells/uL) (HR=0.80 [0.69 - 0.91], p=0.0006) (**Supplementary Figure 2**).

Similar results were seen in an independent cohort of 354 participants (203 [57%] female, 44 [17] years) from the Michigan Genomics Initiative who were treated with the immunosuppressant azathioprine for an auto-immune disease (**Supplementary Table 4**). The baseline WBC count was 8,800 [3,700] cells/uL. The PGS_WBC_ was significantly associated with developing an incident WBC count<3,000 cells/uL after treatment (HR=0.61 [0.38 – 0.99], p=0.04) (**Supplementary Figure 3**).

In sum, a polygenic predisposition toward lower WBC counts was associated with an increased likelihood of having ICD codes for drug toxicity related to a low WBC count and developing leukopenia with treatment.

### Association between the PGS_WBC_ and medication discontinuation

To determine whether the PGS_WBC_ was associated with cessation of medications due to a provider’s concern for a low WBC counts, we examined a cohort of 1,180 BioVU participants (787 [67%] female, 47 [15] years) treated with the azathioprine for an auto-immune disease (**Supplementary Table 5**).^10^ The PGS_WBC_ was inversely associated with baseline WBC count (log difference =0.050 per s.d. change [0.026 - 0.073], p=3.7×10^-5^), but not the change in WBC count during the course of treatment (log change=0.041 [0.27 - 0.350], p=0.79). Azathioprine was discontinued in 34 (3%) participants due to clinical concerns for a low WBC counts The PGS_WBC_ was associated with incident medication discontinuation (HR=0.62 [0.44 - 0.87], p=0.006, adjusted for baseline dose) (**Figure 5**). Thus, a polygenic predisposition toward lower WBC counts was associated with an increased likelihood of stopping an immunosuppressive medication due to concerns for a lower WBC count.

**Figure 5:**
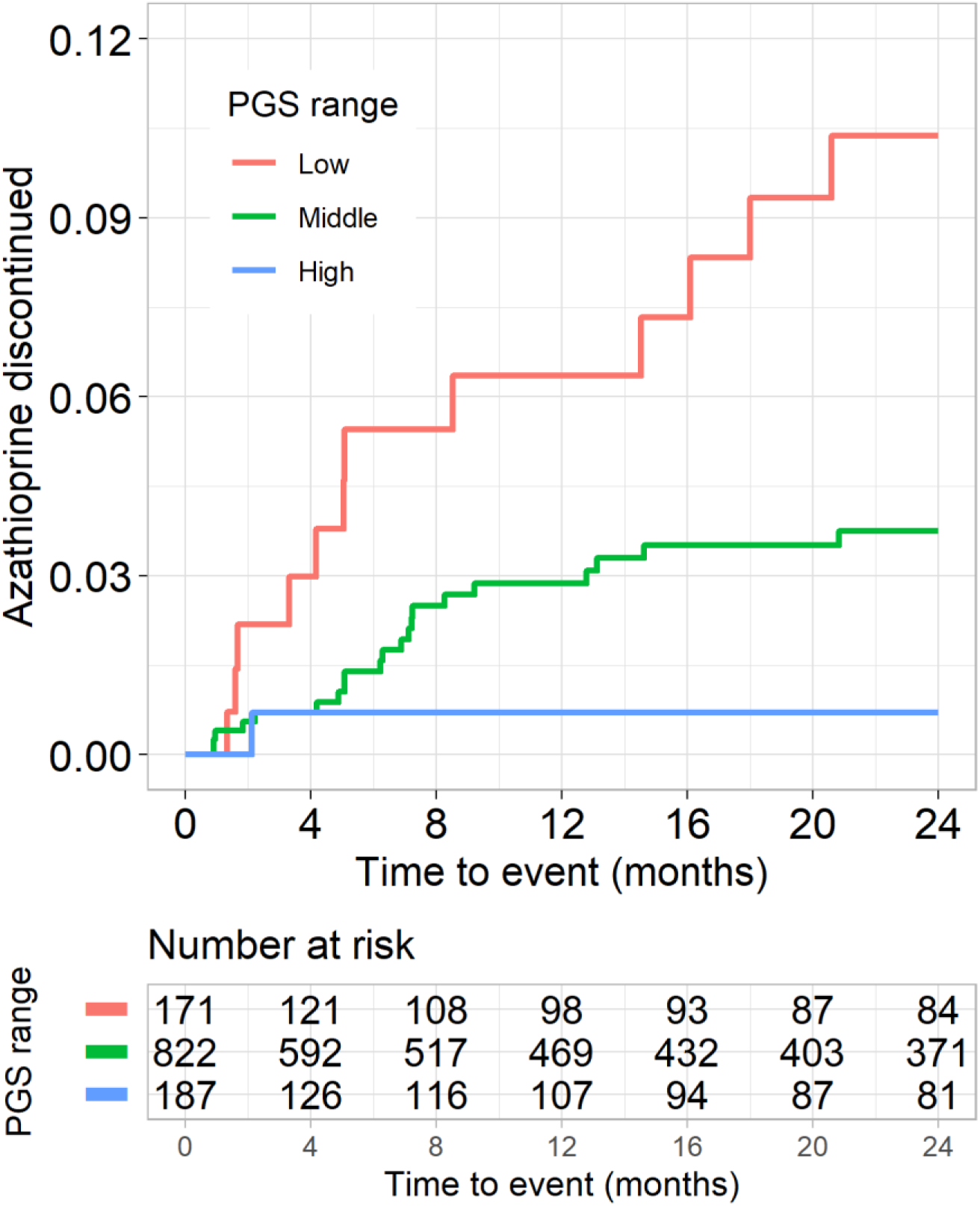
Azathioprine discontinuation. Kaplan-Meier plot for discontinuation of azathioprine due to clinical concern of a low WBC count related to medication toxicity among 1,180 BioVU participants with auto-immune disease. The PGS_WBC_ strata are: Low (<1 s.d. below the mean), Middle (≥ -1 s.d. and ≤ 1 s.d.), High (>1 s.d).

## DISCUSSION

We examined whether a polygenic predisposition to lower WBC counts was associated with clinically significant endpoints. This predisposition was associated with an increased likelihood of having a WBC count falling outside the reference range and receiving an ICD code specifically noting an outlying value. It was also associated with an increased likelihood that a hematologist would list a low WBC count as a reason for performing a bone marrow biopsy, but a decreased likelihood that that such biopsies identified disease within the bone marrow. Finally, a predisposition to lower WBC counts increased the likelihood of drug-induced leukopenia with treatment using chemotherapeutic or immunosuppressant therapies, and increased the likelihood of cessation of azathioprine due to clinical concern for low WBC counts. Collectively, these analyses demonstrate that genetic variation which does not contribute to disease risk, is associated with significant clinical consequences in the form of undergoing diagnostic procedures or altering in therapeutic regimens.

A broad range of biomarkers, including WBC counts, are measured in clinical settings to guide clinical care. Many of these biomarkers are not direct mediators of disease but fluctuate in response to acute and chronic illness. Thus, they are often used to define the characteristics of an illness, which helps establish a differential diagnosis. These biomarkers are paired with clinical references ranges which are used to standardize the delivery of health care. These reference ranges are typically constructed such at 5% of healthy individuals will have a measure that lies outside of the interval and, thus, are outliers.^1,22^ While outlying values are common among healthy individuals predisposed to higher or lower values, they may prompt diagnostic evaluations that often do not identify pathology. This diagnostic uncertainty is stressful to patients, and often leads to serial lab monitoring with no clearly defined endpoint.

The ability of genetic variation associated with benign biomarker variation to systematically disadvantage an identifiable subset of individuals is exemplified by the rs2814778-CC genotype, which is predominantly carried by individuals of African Ancestry and underlies the clinical phenomenon of “benign neutropenia”.^23^ Genotype carriers are predisposed to WBC counts that fall below clinical reference ranges, often because these reference ranges are not derived from individuals who carry the genotype.^5,24^ Consequently, their basal WBC counts are misinterpreted as being inappropriately low, which can lead to clinical investigations that include invasive diagnostic procedures, or to alterations in clinical management including the use of lower medication doses or even discontinuation of medications.^8,10,19,20^ Because this genotype is strongly linked to ancestry, this variant also underlies a measurable racial disparity in clinical medicine.^11,25^

The current study demonstrates that common polygenic variation associated with a diagnostic biomarker can push individuals toward the same clinical endpoints as a genotype with a large effect size. Like WBC counts, many of the biomarkers that are used to guide clinical decision-making have a significant heritable component.^26^ Thus, the implication of these findings is that many individuals are likely predisposed to being identified as outliers for a range of biomarkers because they carry an intrinsic, benign genetic predisposition toward falling outside the reference ranges. As these analyses demonstrate, identifying these individuals based on clinical factors is not always feasible, even by specialists. For instance, the PGS_WBC_ was associated with the clinical indication for a bone marrow biopsy when the indication included a concern for low WBC count expressed by the hematologist.

The pharmacogenomics studies point to the likely mechanism by which this genetic variation drives clinical actions. The PGS_WBC_ was inversely associated with baseline WBC counts in each study, confirming that a genetic predisposition to lower WBC counts has a measurable effect on levels. However, it was not associated with a larger drop in WBC counts with treatment. Collectively, these observations suggest that the reason the PGS_WBC_ associated with outcomes of drug-induced leukopenia and medication cessation is that predisposed individuals have WBC counts that fluctuate across a lower range of values that are close to clinical decision thresholds.

Recognition of the potential harms of ignoring the benign lower WBC counts associated with the rs2814778-CC genotype is prompting changes in practice patterns to mitigate disparities faced by genotype carriers. For instance, there is now an effort to establish treatment guidelines to avoid the inappropriate cessation of medications. Treatment guidelines for the use of clozapine, an antipsychotic which can lower WBC counts, now include provisions that permit lower WBC count discontinuation thresholds for individuals with a diagnosis of benign neutropenia.^27^ However, a limitation of these guidelines is that they often incorporate race as a proxy for genotype, which can be inaccurate.^28^

Similar to the benign neutropenia genotype, a tool like the PGS_WBC_ may provide a means to circumvent futile diagnostic odysseys or unnecessary alterations in clinical care. However, as compared to a discrete genotype, translating benign variation attributable to polygenic risk to the clinical setting is more nuanced, as the exposure is continuous, and a discrete group of individuals cannot be identified. However, possible implementation strategies include the use of clinical reference ranges that are calibrated to an individual’s polygenic predisposition (such as those presented in **Figure 2a**). An alternative strategy is generating prior probabilities, based on a PGS_WBC_, that a given diagnostic work-up (such as a bone marrow biopsy) will identify pathology when performed for an indication of a low WBC count (such as presented in **Figure 3b**).

Finally, these studies identify a novel application of polygenic variation to clinical medicine. There has been considerable interest in using polygenic risk scores (PRS) in clinical settings to identify healthy individuals at risk for future disease.^29,30^ This application of genetics leads to an escalation of clinical care for individuals identified as high-risk, which always entails some degree of risk. A challenge of PRS-based clinical predictors is that they typically have modest discriminative capabilities with respect to developing incident disease.^31,32^ With a modestly discriminative biomarker, the risks will often outweigh the benefits when clinical care is escalated. In contrast to the PRS prediction paradigm, a “benign” PGS_WBC_ could have a role in de-escalating care. For some individuals, a polygenic predictor such as the PGS_WBC_ may provide a means to circumvent otherwise a futile diagnostic odyssey and provide a biologically-motivated explanation for an outlying WBC count. This application of polygenic predictors is apt to have a more favorable risk-benefit ratio, as the default pathway for individuals with outlying values is an escalation in clinical care.

There are strengths to the current study. In particular, it demonstrated a consistent pattern of associations across a diverse range of clinical outcomes and settings. There are also limitations. It is possible that this PGS_WBC_ includes SNPs associated with significant diseases that decrease WBC counts. Mitigating this concern is the fact that the SNP weightings used to construct the PGS_WBC_ were derived from a very large GWAS study where most individuals would not express a given disease; thus, the influences of individuals with overt disease on SNP weightings would be expected to be small. These analyses were restricted to European ancestries, as it has been previously demonstrated that a benign common genetic variant drives these same associations in African ancestries.^9,10^

In conclusion, a polygenic predisposition toward benign lower WBC counts was associated with a diagnosis of leukopenia, undergoing diagnostic procedures and medication discontinuation. Collectively, these studies describe a genetic tool that may have a role in personalizing biomarker WBC count reference ranges for the purpose of de-escalating unnecessary diagnostic investigations or alterations to clinical care.

## ONLINE METHODS

### Study populations

#### BioVU

Study populations were derived from Vanderbilt University Medical Center’s (VUMC) DNA biobank resource (BioVU). BioVU comprises 270,000 consented participants and is constructed from discarded blood samples collected from consented individuals and linked to de-identified electronic health records (EHR).^33^ The de-identified EHR (the “synthetic derivative”) captures a large portion of data available through the medical center’s electronic health record.^33^ Analyses were restricted to 71,078 participants of White European Ancestry with existing SNP genotyping. All BioVU studies were evaluated by the VUMC Institutional Review Board (IRB) and determined to be non-human subjects research. The following cohorts were derived from this population:

1. WBC cohort: This cohort of 11,694 BioVU participants was used to define the relationship between the white blood cell count PGS and measured white cell counts. To minimize the likelihood that WBC counts were collected during an acute illness, the population was restricted to individuals with one or more WBC counts measured contemporaneously with an International Classification of Disease^34^ (ICD)-9/10 code for a routine health exam (ICD-9: V70.9, V20, V20.1, V70, V70.0, V20.2; ICD-10 Z00.8, Z00.129, Z00.00, Z00.01, Z00.121). These codes are typically used in a primary care setting to denote a well-visit encounter. Participants with an ICD code for a hematological malignancy, chemotherapy or radiation therapy (ICD codes listed in **Supplementary Materials**), as these diagnoses or treatments can markedly impact WBC counts.
2. Bone marrow biopsy cohort: Derivation of this cohort followed previously described methods.^12^ Briefly, among 2,302 BioVU participants with a biopsy pathology report in their clinical record, participants were excluded if they had an ICD code for hematological malignancy, blood transfusion, organ transplant, or chemotherapy/radiation prior to their first biopsy. There were 922 participants after these exclusions. From this set, participants were further excluded if there was not a text phrase pertaining to a low WBC count in the clinical indication portion of the biopsy report (text phrases are listed in **Supplementary Table 6**). Participants were further excluded if, upon manual review of their first available biopsy report, they were noted to have an established hematological cancer or hematological diagnosis. After exclusions, there were 117 participants in the final cohort without a known diagnosis of malignancy who underwent a first biopsy for an indication that included a WBC count.
3. Taxane cohort: Participants were from a previously curated multi-ancestry longitudinal cohort of 3,492 BioVU participants undergoing treatment for a primary cancer with taxane chemotherapies (paclitaxel or docetaxel).^21^ The cohort was constructed to examine racial differences in rates of incident drug-induced neutropenia among participants of black (n=365) and white (n=3019) races. Analyses were restricted to 1,724 European ancestry participants with existing genotyping and a baseline white blood cell (n=1,721) or neutrophil count (n=1,724) >1000 cells/µL.
4. Azathioprine cohort: Participants were from a previously curated multi-ancestry longitudinal cohort of 1,466 BioVU participants with auto-immune or other inflammatory diseases who were newly started on the drug azathioprine.^10^ The cohort was constructed to identify genetic factors associated with incident discontinuation of azathioprine among participants of black (n=165) and white races (n=1,301). The analyses were restricted to the subset of 1,180 European ancestry participants ages 20-80 years.

#### eMERGE cohort

The eMERGE consortium is a collection of institutions with DNA biobanks that are linked to electronic health records. These analyses examined adults of White European ancestry born before 1990 from the eMERGE network (phases 1-3)^35^. The participating eMERGE sites were Columbia University, Geisinger, Marshfield Clinic, Northwestern University, Mayo Clinic, Harvard University, Mt. Sinai Health System, and Kaiser Permanente/University of Washington, Seattle. These analyses were approved by each eMERGE institution’s IRB.^35^ Participants with an ICD code for a hematological malignancy, chemotherapy or radiation therapy (ICD codes listed in **Supplementary Materials**) were excluded from the analyses. After exclusions, there were 18,218 (n=10,162 [56%] female) adult participants of European ancestry.

Michigan Genomics Initiative (MGI) cohort: Study participants are derived from a biobank of patients recruited though the Michigan Medicine health system.^36^ As of 2/2023, approximately 91,000 patients have consented to the linkage of a DNA sample with their Michigan Medicine electronic health record for research purposes. This analysis was performed on the “Freeze 5” dataset of MGI (n=70,266), and restricted to 59,910 samples of European ancestry based on genetic principal component analysis. Participants were selected for inclusion if they had an ICD code for inflammatory disease (inflammatory bowel disease or connective tissue diseases) and had azathioprine on their medication list subsequent to receiving the ICD code. Participants were excluded if they had an ICD code for solid organ transplant, a hematological malignancy or a stem cell transplant. There were 417 participants after these exclusions. Participants were further excluded if they did not have a WBC count measure taken after azathioprine initiation. The final set comprised 354 participants.

### Genetic data

BioVU participants were genotyped on the Illumina Infinium MEGA^EX^ platform. SNP genotyping was called by the Vanderbilt Technologies for Advanced Genomics Analysis and Research (VANTAGE) Design core.^37,38^ Genetic ancestry was determined by principal components (PCs) analysis in conjunction with HapMap reference populations and individuals who did not fall within with 4 standard deviations of White European populations were excluded. Participants were further excluded for outlying heterozygosity measures, a discordant genetically-determined vs. reported sex, or >4% missing genetic data. Prior to imputation, data were put through the HRC-1000G check tool (v4.2.5) and pre-phased using Eagle v2.4.1.^39^ QC analyses were performed using PLINK v1.90b6.17 and v2.00a3LM.^40^ Imputation was performed using the Michigan Imputation Server in conjunction and the HRC v1.1 reference panel.^41^ Imputed data were filtered for a sample missingness rate <2%, a SNP missingness rate <4%, and SNP deviation from Hardy-Weinberg p<10^-6^. PCs of ancestry were calculated across the entire BioVU cohort using the SNPRelate package.^42^

eMERGE participants were genotyped on multiple platforms and underwent a similar QC analysis and imputation protocol as BioVU participants, as previously described.^43,44^

MGI samples are genotyped in waves based on time of recruitment, with initial waves genotyped on a customized Illumina Infinium CoreExome genotyping array and subsequent waves on a customized Illumina Infinium Global Screening Array. Genotypes are then imputed to 307,883,040 variants using the Trans-Omics for Precision Medicine (TOPMed) haplotype reference panel. 50,463,429 variants passed standard post-imputation filters, which removed poorly imputed variants with Rsq < 0.3 and very rare variants with minor allele frequency (MAF) < 0.01%.

### Development of a benign WBC polygenic score (PGS_WBC_)

The polygenic score (PGS) was derived from WBC count summary statistics from the European Ancestry subset of a genome-wide association study (GWAS) of hematological traits among ∼750,000 participants.^15^ A LD-reduced set (r-square<0.01) of non-palindromic SNPs associated with WBC count (p<5×10^-6^, minor allele frequency=0.01, imputation r-square≥0.7) was selected using a clumping algorithm.^45^ The p-value threshold was selected because it had the highest linear partial correlation (r=0.31, adjusted for age and sex) between the PGS and measured WBC counts among three thresholds examined (p<5×10^-8^, 5×10^-7^, 5×10^-6^) in a set of 11,694 participants from BioVU without underlying hematological disease (the “WBC Cohort” described above). SNPs located in the Major Histocompatibility Complex genomic region (6:25500000-33500000), which is associated with multiple auto-immune diseases, were excluded. To identify SNPs associated with hematological diseases or other diseases that cause low WBC counts, all GWAS SNPs with a WBC count association p<5×10^-6^ were interrogated against associations curated in the GWAS Catalog,^46^ and those SNPs that were associated with other phenotypes at p<5×10^-7^ were identified. There were 1,507 GWAS Catalog phenotypes associated with 1 or more SNPs. Among these were 21 hematological malignancies and systemic lupus erythematous, all of which are important diagnoses on the differential diagnosis for a low WBC count (see list in **Supplementary Table 7**).^17^ All SNPs in the PGS_WBC_ that were in LD (r-square>0.5) with a SNP associated with these 22 diagnoses were excluded. After exclusions, there were 1,739 WBC-associated SNPs used to compute the PGS_WBC_ (a listing of SNPs and weights can be found in **Supplementary Materials**). A weighted PGS_WBC_ was calculated for each participant by summing the product of the allele dosage and the SNP weighting from the WBCs GWAS for each SNP.

### Measured white blood cell counts

For the WBC cohort, WBC counts collected on the same day as a routine health visit were extracted. WBC counts >35,000 cells/µL were excluded, as these values likely represent an active disease process. Each WBC count measure had an associated reference range that was specific to the clinical assay, with a lower bound typically of ∼3,900 cells/µL. A participant was defined as an outlier if they had a WBC count measure below the lower bound indicated by the assay.

### ICD code based phenotypes

Phecodes are collections of related ICD-9/ICD-10 diagnosis codes (Phecodes definitions can be found at https://phewas.mc.vanderbilt.edu/).^47,48^ For each Phecode, cases are participants with one or more instances of the relevant ICD codes appearing in their medical record. Controls are participants without those codes and whose age fell within the range of ages observed among cases. The Phecodes codes examined were for a low WBC count and low neutrophil count (neutropenia), and codes related to hematological malignancies (n=15) and auto-immune diseases (n=22) that were prevalent (n>100 cases) among the BioVU participants. The list of Phecodes can be found in **Supplementary Table 1**.

For the phenotype of toxicity related to antineoplastic and immunosuppressive medications, cases were participants having any of the following ICD-9 (963.1, 960.7, 284.11, E933.1, E930.7) or ICD-10 (T45.1X4D, T45.1X4S, T45.1X5D, T45.1X5S, T45.1X5A, T45.1X1S, T45.1X4A, T45.1X2A, T45.1X1A) codes. To evaluate the specificity of the association for a low WBC count, cases were dichotomized into those with and those without a clinical note containing a mention of a low WBC count (keywords are listed in **Supplementary Table 8**) entered on the same day as the toxicity code.

### Bone marrow biopsy phenotypes

Among 922 BioVU participants with a bone marrow biopsy report in their clinical record and without a prior history of a hematological cancer or chemotherapy, there were 117 participants biopsied for a low WBC count (the Biopsy Cohort). The first biopsy report was reviewed, and extracted data included hematological comorbidities (related to platelets, red blood cell counts) and other comorbidities listed in the clinical history. The primary outcome was a determination by the pathologist as to whether a clinically significant marrow abnormality was present (coded as “normal” or “abnormal”).^12^ Data were extracted by a physician (JDM) and hematologist (SCB) to a REDCap database.

### Pharmacogenomic phenotypes

For the taxane longitudinal study, the primary outcome was development of a WBC count<3,000 cells/µL during the first cycle of treatment. Participants were censored at the earlier of a primary outcome event, the start of their second cycle of chemotherapy, or 1 month after the initiation of treatment. Secondary analyses were performed the examine an incident outcome of absolute neutrophil count <1,500 cells/µL.

For the MGI azathioprine study, the primary outcome was an incident WBC count<3,000 cells/µL. In secondary analyses, thresholds of <3,500 and <4,000 cells/µL were also examined to demonstrate that associations were consistent when using higher thresholds where larger numbers of participants met the threshold criteria. Participants were censored from the study at the earlier of a primary outcome event, or their last clinical encounter up to 24 months. The PGS_WBC_ comprised a subset of 1,704 SNPs that passed quality control.

For the BioVU azathioprine discontinuation study, the primary outcome was azathioprine discontinuation due to leukopenia or neutropenia, based on a provider’s assessment, within 24 months of drug initiation. All participants were censored from the study at the earlier of a primary outcome event, time of drug discontinuation, or their last clinical encounter up to 24 months. In this separately genotyped population, the PGS_WBC_ comprised a subset of 1,680 SNPs that passed quality control.

### Analysis

The PGS_WBC_ was mean standardized, and association statistics reflect changes per standard deviation (s.d.) increase in the PGS_WBC_. A lower value reflects a polygenic predisposition toward lower WBC counts, and an inverse association between the PGS_WBC_ and an outcome indicates that a predisposition to lower counts increases the likelihood of the outcome.

In the full BioVU cohort, multivariable logistic regression was used to determine whether the PGS_WBC_ was associated with having a pheWAS code-based diagnosis of 15 bone marrow malignancies and 22 rheumatological diseases prevalent in this cohort. Models were adjusted age, sex, and 5 PCs. To account for multiple testing, and FDR p<0.05 was considered significant.

WBC cohort: The observed distribution of WBC counts across the range of PGS_WBC_ values was visualized by computing the 5^th^, 50^th^ and 95^th^ percentiles of WBC counts within sequential windows (+/- 0.2 s.d.) across the range of the PGS_WBC_. Multivariable logistic regression, adjusting for age, age-squared, sex, and 5 PCs was used to test the association between the PGS_WBC_ and (1) having a WBC count that fell outside the reference range and (2) having a Phecode for a low WBC count (phecode 288.1). The association with the Phecode was also tested in the eMERGE cohort.

Multivariable logistic regression, adjusting age, sex, and 5 PCs, was used to determine whether the PGS_WBC_ was associated with having a bone marrow biopsy for a clinical indication of low WBC count, among the 922 participants with a bone marrow biopsy.

Bone marrow biopsy cohort: Multivariable logistic regression was used to test the association between the PGS_WBC_ and having a normal bone marrow biopsy finding, adjusting for age, sex, 5 PCs, and hematological comorbidities (low platelets, low red cells, other hematological abnormalities).

In the full BioVU cohort, multivariable logistic regression was used to determine whether the PGS_WBC_ was associated an ICD-based diagnosis of drug toxicity, with or without mention of a low WBC count. Models were adjusted age, sex, and 5 PCs.

Taxane and Azathioprine cohorts: Multivariable linear regression was used to determine the association between PGS_WBC_ and baseline log-transformed WBC count and change in WBC count (count at baseline - count at censoring time). A Cox proportional hazards regression model was used to measure the hazard ratio associated with either incident leukopenia/neutropenia (taxanes) or azathioprine discontinuation for low counts. All analyses were adjusted for sex, age at drug initiation, and either 5 (taxane and azathioprine) or 10 (azathioprine discontinuation) PCs. The fit of the Cox models was assessed using analyses of Schoenfeld (proportional hazards assumption), Martingale (non-linearity) and deviance residuals (outliers). Data were visualized by Kaplan-Meier analysis.

All statistical tests were two-sided and a p<0.05 was considered significant unless otherwise noted. Statistical analyses were conducted using R 4.2.0.

## DATA AVAILABILITY

Subject-level access to BioVU clinical and genetic data is controlled by the BioVU data repository (https://victr.vumc.org/biovu-description/#). Upon publication, data sets of individual-level phenotype data and corresponding data dictionaries to replicate the primary findings presented here for research purposes will be made available upon request from the repository. BioVU vetting for use of individual-level data includes institutional IRB approval, data use agreements, and administrative and scientific reviews.

## CODE AVAILABILITY

No custom computer code or algorithms were used to generate results that are reported in the paper.

## AUTHOR CONTRIBUTIONS

JDM had full access to all data in the trial and takes responsibility for the integrity of the data and the accuracy of the data analysis. JDM, ALD, JZ, NSZ, CMS, SLV, SCB, XS, CMC and VKK provided substantial contributions to the design of the study. LD, NSZ, LB, WQW, MS, GPJ, EAR, AK, AS, IJK, TLW, JG, HH, NJC, DMR, SCB, XO, SGF, BV and CPC made substantial contributions to the acquisition of data. JDM performed the primary analysis of the BioVU data. MZ performed analyses of the MGI data. JDM, CMS, JPS and VKK wrote the first draft of the manuscript. All coauthors critically reviewed the manuscript for important intellectual content, provided final approval of the version to be published, and agree to be accountable for all aspects of the work presented.

## ACKNOWLEDGEMENTS

The authors acknowledge the MGI participants, Precision Health at the UM, the UM Medical School Central Biorepository, and the UM Advanced Genomics Core for providing data and specimen storage, management, processing, and distribution services and the Center for Statistical Genetics in the Department of Biostatistics at the School of Public Health for genotype data curation, imputation, and management in support of the research reported in this publication.

## REFERENCES

1. Ozarda, Y. Reference intervals: current status, recent developments and future considerations. Biochem Med (Zagreb) 26, 5–16 (2016).

2. Higgins, V., Nieuwesteeg, M. & Adeli, K. Chapter 3 - Reference intervals: theory and practice. in Contemporary Practice in Clinical Chemistry (Fourth Edition) (eds. Clarke, W. & Marzinke, M. A.) 37–56 (Academic Press, 2020). doi:10.1016/B978-0-12-815499-1.00003-X.

3. Valent, P. Low blood counts: immune mediated, idiopathic, or myelodysplasia. Hematology Am Soc Hematol Educ Program 2012, 485–491 (2012).

4. Boxer, L. A. How to approach neutropenia. Hematology Am Soc Hematol Educ Program 2012, 174–182 (2012).

5. Reich, D. et al. Reduced neutrophil count in people of African descent is due to a regulatory variant in the Duffy antigen receptor for chemokines gene. PLoS Genet 5, e1000360– e1000360 (2009).

6. Reiner, A. P. et al. Genome-wide association study of white blood cell count in 16,388 African Americans: the continental origins and genetic epidemiology network (COGENT). PLoS Genet 7, e1002108 (2011).

7. Manu, P., Sarvaiya, N., Rogozea, L. M., Kane, J. M. & Correll, C. U. Benign Ethnic Neutropenia and Clozapine Use: A Systematic Review of the Evidence and Treatment Recommendations. J Clin Psychiatry 77, e909–916 (2016).

8. Hershman, D. et al. Ethnic neutropenia and treatment delay in African American women undergoing chemotherapy for early-stage breast cancer. J Natl Cancer Inst 95, 1545–1548 (2003).

9. Van Driest, S. L. et al. Association Between a Common, Benign Genotype and Unnecessary Bone Marrow Biopsies Among African American Patients. JAMA Internal Medicine 181, 1100–1105 (2021).

10. Dickson, A. L. et al. Race, Genotype, and Azathioprine Discontinuation: A Cohort Study. Ann Intern Med (2022) doi:10.7326/M21-4675.

11. Vastola, M. E. et al. Laboratory Eligibility Criteria as Potential Barriers to Participation by Black Men in Prostate Cancer Clinical Trials. JAMA Oncol 4, 413–414 (2018).

12. Borinstein, S. C. et al. Frequency of benign neutropenia among Black versus White individuals undergoing a bone marrow assessment. J Cell Mol Med 26, 3628–3635 (2022).

13. Astle, W. J. et al. The Allelic Landscape of Human Blood Cell Trait Variation and Links to Common Complex Disease. Cell 167, 1415–1429.e19 (2016).

14. Keller, M. F. et al. Trans-ethnic meta-analysis of white blood cell phenotypes. Hum. Mol. Genet. 23, 6944–6960 (2014).

15. Chen, M.-H. et al. Trans-ethnic and Ancestry-Specific Blood-Cell Genetics in 746,667 Individuals from 5 Global Populations. Cell 182, 1198–1213.e14 (2020).

16. Karnes, J. H. et al. Phenome-wide scanning identifies multiple diseases and disease severity phenotypes associated with HLA variants. Sci Transl Med 9, (2017).

17. Kandane-Rathnayake, R. et al. Independent associations of lymphopenia and neutropenia in patients with systemic lupus erythematosus: a longitudinal, multinational study. Rheumatology (Oxford) 60, 5185–5193 (2021).

18. Israel, R. A. The International Classification of Disease. Two hundred years of development. Public Health Rep 93, 150–152 (1978).

19. Van Driest, S. L. et al. Association Between a Common, Benign Genotype and Unnecessary Bone Marrow Biopsies Among African American Patients. JAMA Intern Med (2021) doi:10.1001/jamainternmed.2021.3108.

20. Legge, S. E. et al. A genome-wide association study in individuals of African ancestry reveals the importance of the Duffy-null genotype in the assessment of clozapine-related neutropenia. Mol Psychiatry 24, 328–337 (2019).

21. Zheng, N. S. et al. Racial disparity in taxane-induced neutropenia among cancer patients. Cancer Med 10, 6767–6776 (2021).

22. Rappoport, N. et al. Comparing Ethnicity-Specific Reference Intervals for Clinical Laboratory Tests from EHR Data. J Appl Lab Med 3, 366–377 (2018).

23. Tournamille, C., Colin, Y., Cartron, J. P. & Le Van Kim, C. Disruption of a GATA motif in the Duffy gene promoter abolishes erythroid gene expression in Duffy-negative individuals. Nat. Genet. 10, 224–228 (1995).

24. Nalls, M. A. et al. Admixture mapping of white cell count: genetic locus responsible for lower white blood cell count in the Health ABC and Jackson Heart studies. Am. J. Hum. Genet. 82, 81–87 (2008).

25. Merz, L. E. & Achebe, M. When non-Whiteness becomes a condition. Blood 137, 13–15 (2021).

26. Enroth, S., Johansson, A., Enroth, S. B. & Gyllensten, U. Strong effects of genetic and lifestyle factors on biomarker variation and use of personalized cutoffs. Nat Commun 5, 4684 (2014).

27. FDA. FDA Drug Safety Communication: FDA modifies monitoring for neutropenia associated with schizophrenia medicine clozapine; approves new shared REMS program for all clozapine medicines. (2015).

28. Sirugo, G. & Wonkam, A. Beyond Race: A Wake-up Call for Drug Therapy Informed by Genotyping. Ann Intern Med 175, 1187–1188 (2022).

29. Torkamani, A., Wineinger, N. E. & Topol, E. J. The personal and clinical utility of polygenic risk scores. Nat. Rev. Genet. 19, 581–590 (2018).

30. Khera, A. V. et al. Genome-wide polygenic scores for common diseases identify individuals with risk equivalent to monogenic mutations. Nat. Genet. (2018) doi:10.1038/s41588-018-0183-z.

31. Wald, N. J. & Old, R. The illusion of polygenic disease risk prediction. Genet. Med. 21, 1705–1707 (2019).

32. Sud, A., Turnbull, C. & Houlston, R. Will polygenic risk scores for cancer ever be clinically useful? NPJ Precis Oncol 5, 40 (2021).

33. Roden, D. M. et al. Development of a large-scale de-identified DNA biobank to enable personalized medicine. Clin. Pharmacol. Ther. 84, 362–369 (2008).

34. Denny, J. C. et al. PheWAS: demonstrating the feasibility of a phenome-wide scan to discover gene-disease associations. Bioinformatics 26, 1205–1210 (2010).

35. Gottesman, O. et al. The Electronic Medical Records and Genomics (eMERGE) Network: past, present, and future. Genet. Med. 15, 761–771 (2013).

36. Zawistowski, M. et al. The Michigan Genomics Initiative: A biobank linking genotypes and electronic clinical records in Michigan Medicine patients. Cell Genom 3, 100257 (2023).

37. Guo, Y. et al. Illumina human exome genotyping array clustering and quality control. Nat Protoc 9, 2643–2662 (2014).

38. Mosley, J. D. et al. Identifying genetically driven clinical phenotypes using linear mixed models. Nat Commun 7, 11433 (2016).

39. Pasaniuc, B. et al. Fast and accurate imputation of summary statistics enhances evidence of functional enrichment. Bioinformatics 30, 2906–2914 (2014).

40. Purcell, S. et al. PLINK: a tool set for whole-genome association and population-based linkage analyses. Am. J. Hum. Genet. 81, 559–575 (2007).

41. McCarthy, S. et al. A reference panel of 64,976 haplotypes for genotype imputation. Nat. Genet. 48, 1279–1283 (2016).

42. Zheng, X. et al. A high-performance computing toolset for relatedness and principal component analysis of SNP data. Bioinformatics 28, 3326–3328 (2012).

43. Zuvich, R. L. et al. Pitfalls of merging GWAS data: lessons learned in the eMERGE network and quality control procedures to maintain high data quality. Genet. Epidemiol. 35, 887–898 (2011).

44. Stanaway, I. B. et al. The eMERGE genotype set of 83,717 subjects imputed to ∼40 million variants genome wide and association with the herpes zoster medical record phenotype. Genet. Epidemiol. (2018) doi:10.1002/gepi.22167.

45. International Schizophrenia Consortium et al. Common polygenic variation contributes to risk of schizophrenia and bipolar disorder. Nature 460, 748–752 (2009).

46. Buniello, A. et al. The NHGRI-EBI GWAS Catalog of published genome-wide association studies, targeted arrays and summary statistics 2019. Nucleic Acids Res. (2018) doi:10.1093/nar/gky1120.

47. Wei, W.-Q. et al. Evaluating phecodes, clinical classification software, and ICD-9-CM codes for phenome-wide association studies in the electronic health record. PLoS ONE 12, e0175508 (2017).

48. Wu, P. et al. Mapping ICD-10 and ICD-10-CM Codes to Phecodes: Workflow Development and Initial Evaluation. JMIR Med Inform 7, e14325 (2019).

